# Outcomes evaluated in controlled clinical trials on the management of COVID-19: A methodological systematic review

**DOI:** 10.1101/2020.10.26.20218370

**Authors:** Alexander G. Mathioudakis, Markus Fally, Rola Hashad, Ahmed Kouta, Ali Sina Hadi, Sean Blandin Knight, Nawar Diar Bakerly, Dave Singh, Paula R. Williamson, Timothy Felton, Jørgen Vestbo

**Author notes:** **Corresponding Author:** Prof. Jørgen Vestbo DMSc, FRCP, FERS, FMedSci, Professor of Respiratory Medicine, Division of Infection, Immunity and Respiratory Medicine, School of Biological Sciences, The University of Manchester, UK. AGM and MF contributed equally to this work. **Author contribution:** Study conception: AGM, MF, TF and JV. Study design: AGM, MF, TF and JV. Data collection: AGM, MF, RH, AK, ASH. Data analysis: AGM. Methodological expertise: AGM, MF, PRW, JV. Interpretation of the findings: All authors. Manuscript preparation: AGM. Critical revision of the manuscript: All authors. Final approval of the manuscript: All authors.

## Abstract

It is crucial that randomized controlled trials (RCTs) on the management of coronavirus disease 2019 (COVID-19) evaluate the outcomes that are critical to patients and clinicians, to facilitate relevance, interpretability, and comparability.

This methodological systematic review describes the outcomes evaluated in 415 RCTs on the management of COVID-19, that were registered with ClinicalTrials.gov, by 5/5/2020.

Significant heterogeneity was observed in the selection of outcomes and the instruments used to measure them. Mortality, adverse events and treatment success or failure are only evaluated in 64.4%, 48.4% and 43% of the included studies, respectively, while other outcomes are selected less often. Studies focusing on more severe presentations (hospitalized patients or requiring intensive care) most frequently evaluate mortality and adverse events, while hospital admission and viral detection/load are most frequently assessed in the community setting. Outcome measurement instruments are poorly reported and heterogeneous. In general, simple instruments that can control for important sources of bias are favoured. Follow-up does not exceed one month in 64.3% of these earlier trials, and long-term COVID-19 burden is rarely assessed.

The methodological issues identified could delay the introduction of potentially life-saving treatments in clinical practice. Our findings demonstrate the need for consensus in the design of RCTs.

**Take home message:** @ERSpublications: This systematic review describes the heterogeneity in outcomes evaluated in 415 RCTs on COVID-19 management and the instruments used to measure them. Our findings reveal a need for consensus in the design of future RCTs.

## Background

Within a few months from its emergence, the Coronavirus Disease 2019 (COVID-19), characterized by an excessive infectivity, morbidity and mortality, evolved into a global, lethal pandemic^1^. Faced with the risk of a public health disaster, the biomedical community sparked an unprecedented research mobilization aiming to understand the virus and develop effective preventive and therapeutic strategies^2^. Characteristically, within ten months, over 60 thousand publications focusing on COVID-19 were indexed in the PubMed database and almost two thousand interventional studies were registered with the ClinicalTrials.gov database.

However, inevitably, the limited knowledge about the disease and the need for an expeditious response to the unfolding pandemic did not allow, in some cases, for adequate methodological planning and co-ordination. Firstly, extensive research duplication (or -better-multiplication) has been observed, with numerous randomized controlled trials (RCTs) evaluating the same interventions for COVID-19 in parallel^3^. This fragmentation of the research data could delay the acquisition of confident, actionable results regarding each treatment. Moreover, lack of standardization in trial design could limit comparability. An important source of variability in trial design could arise from the outcomes (endpoints) that are selected for evaluation. Heterogeneity in trial outcomes and omission of outcomes that are critically important to patients and clinicians complicate interpreting, comparing and synthesizing trial results, potentially delaying the introduction of novel, life-saving treatments into clinical practice^4,5^.

Core outcome sets are developed to address heterogeneity in the selection of outcomes. These are agreed standardized sets of outcomes that should be measured and reported as a minimum in all clinical trials in specific areas of health or health care^6^. The Core Outcome Measures in Effectiveness Trials (COMET) has developed a rigorous methodology for their development, to ensure the most pertinent clinical outcomes are included in core outcome sets^6-8^. COMET recommends the conduct of extensive methodological systematic reviews and qualitative research aiming to develop a longlist of all relevant outcomes for a disease area. This should be followed by a multi-stakeholder Delphi survey, aimed to prioritize the most pertinent outcomes for inclusion in the core outcome set^6-8^.

Upon the emergence of COVID-19 pandemic, there was an urgent need for the development of a core outcome set. Within a few months, four core outcome sets were developed, using an accelerated process^9-12^. These were based on methodological systematic reviews of the first registered RCTs, which were limited in number, but also in design, due to the limited knowledge of the nature of COVID-19, at the time. However, in the meantime, our knowledge of the natural history of COVID-19 is expanding rapidly and numerous clinical trials have been registered in clinical trial registries such as the ClinicalTrials.gov database. In this methodological survey, we describe the outcomes that are tested in RCTs evaluating therapeutic interventions for COVID-19 and the instruments used to measure these outcomes.

## Methods

For the conduct of this methodological systematic review, we followed standard methodology recommended by the Core Outcome Measures in Effectiveness Trials (COMET) initiative^6^, that was successfully applied in previous, similar methodological surveys^13-15^.

### Study selection and data extraction

Planned, ongoing or completed interventional clinical trials evaluating pharmacological or non-pharmacological interventions for the management of COVID-19 were considered eligible. Phase 1 trials were considered beyond the scope of this manuscript and, thus, excluded. All eligible trials from the U.S. National Library of Medicine clinical trials register (ClinicalTrials.gov, searched on May 5^th^, 2020) were retrieved using standard filters recommended by the library. More specifically, for identifying studies evaluating COVID-19, we used the following terms: COVID-19, SARS-CoV-2, severe acute respiratory syndrome coronavirus 2, 2019-nCoV, 2019 novel coronavirus, and Wuhan coronavirus. Only studies identified as interventional by the submitting researcher were retrieved.

Eligible studies were grouped into phase 2 or later stage trials, as we anticipated between-group differences in the selection of outcomes. In addition, trials are presented grouped according to the recruitment setting (community, hospital, or intensive care unit), which can be used as a surrogate measure of the severity of the participants recruited. The main methodological characteristics of all eligible studies were extracted automatically from the ClinicalTrials.gov extract (.csv), using a script developed in the platform R statistics (version 3.4.3; R Foundation for Statistical Computing, Vienna, Austria). One researcher (amongst MF, RH, ASH, AK) confirmed eligibility, cross-checked pre-extracted data for accuracy, searched for additional reports of the study protocol and extracted additional data, that were not automatically captured. A second researcher (AGM) cross-checked all extracted data for accuracy. Disagreement was resolved through discussion. Extracted data included the projected recruitment sizes, study settings, as well as details on the eligibility criteria and evaluated outcome measures.

### Outcome grouping and classification

Detailed descriptions of all outcome measures were extracted verbatim from the study protocols or registry entries. After in-depth assessment of the outcomes evaluated in a random sample of 20 studies, we developed a list of generic outcome categories defined by the treatment effect they aim to capture, rather than the specific measurement instrument. For example, we chose the term “treatment success” to encapsulate outcomes such as the proportion of patients who were completely asymptomatic by a specific timepoint or those who experienced a clinical improvement of at least two points on the 9-category ordinal scale developed by the World Health Organisation (WHO) for COVID-19^16^, by a specific follow-up timepoint. Next, two authors (amongst MF, RH, ASH, AK) categorized each of the extracted outcomes within the generic outcome categories. New generic outcome categories were developed as needed, in cases where the evaluated outcomes did not fit any of the existing categories, based on consensus among the co-authors. The same investigators also captured the instruments used for the quantification of each outcome. Disagreement was resolved through discussion with another reviewer (AGM).

Finally, the generic outcomes were further classified according to the COMET taxonomy^17^.

## Results

### Description of the included studies

Our search retrieved 745 interventional studies. After excluding diagnostic, prognostic, preventive studies, phase 1 trials and those not directly focusing on the management of COVID-19, we selected 415 studies for inclusion in this systematic survey, including 178 phase 2, and 237 later phase RCTs. Details on the selection process (PRISMA flowchart, supplementary figure 1), as well as the trial registration numbers, and planned study population of all included studies are available in the online appendix.

**Figure 1.**
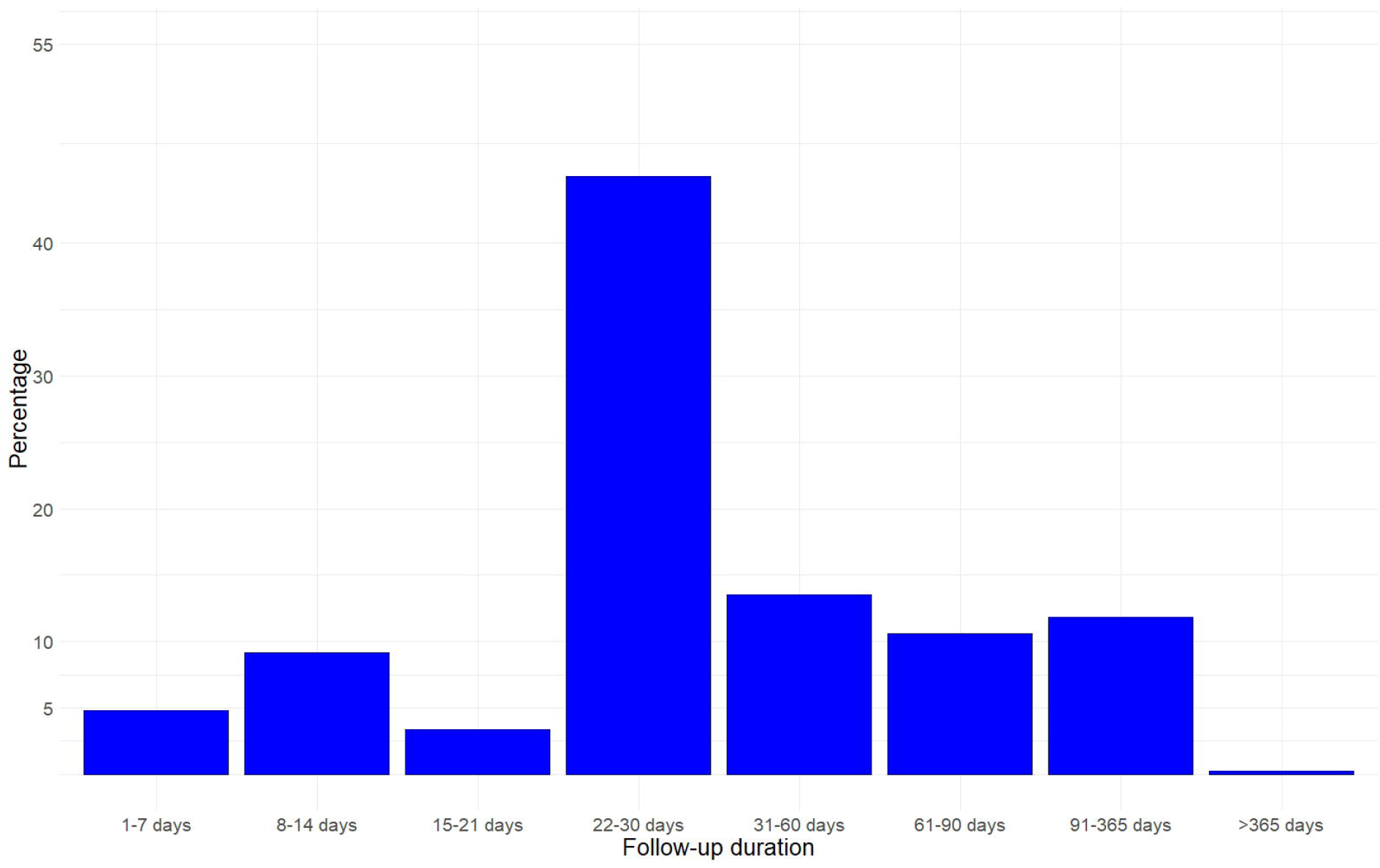
Duration of follow-up in the included studies.

Most of the included trials are conducted by academic investigators (75.7%) and only one in four is sponsored by the pharmaceutical industry. The planned recruitment ranges between 7 and 12,000 participants (median: 160, interquartile range [IQR]: 67-400). Most trials include two intervention arms (74.8%), but one in four evaluates more than two, and up to 19 interventions. Moreover, 79.8% of the trials are conducted in a hospital setting, including 6.5% conducted in the intensive care unit (ICU), while 15.2% are conducted in the community. Descriptions of disease severity are heterogeneous with the recruitment setting being the most consistent measure of disease. Details on the characteristics of the included studies are available in table 1.

**Table 1.** Characteristics of the included studies. *Studies conducted in multiple continents are counted in each participating continent.

|  | Phase 2 trials (n=178) | Later phase trials (n=237) |
| --- | --- | --- |
| **Number of participants** |  |  |
| *Median (range)* | 120 (15-2,000) | 253 (7-12,000) |
| **Setting** |  |  |
| Community | 25 (14.0%) | 38 (16.0%) |
| Hospital | 137 (77.0%) | 167 (70.5%) |
| Community & Hospital | 3 (1.7%) | 1 (0.4%) |
| ICU | 9 (5.1%) | 18 (7.6%) |
| Other | 0 (0.0%) | 1 (0.4%) |
| Unclear | 4 (2.2%) | 13 (5.5%) |
| **Continent** |  |  |
| Africa | 5 (2.8%) | 21 (8.9%) |
| Asia | 29 (16.3%) | 51 (21.5%) |
| Europe | 46 (25.8%) | 94 (39.7%) |
| North America | 90 (50.6%) | 67 (28.3%) |
| Oceania | 1 (0.6%) | 1 (0.4%) |
| South America | 12 (6.7%) | 22 (9.3%) |
| Multiple continents* | 6 (3.4%) | 15 (6.3%) |
| Unclear | 6 (3.4%) | 0 (0.0%) |
| **Age range** |  |  |
| Minimum age, |  |  |
| *Median (range)* | 18 (3-50) | 18 (1-70) |
| *Not reported* | 2 (1.1%) | 0 (0.0%) |
| Maximum age, |  |  |
| *Median (range)* | 80 (50-110) | 80 (40-110) |
| *Not reported* | 115 (64.6%) | 157 (66.0%) |
| **Number of interventions** |  |  |
| 2 | 139 (78.1%) | 172 (72.6%) |
| 3 | 25 (14.0%) | 40 (16.9%) |
| 4 | 10 (5.6%) | 11 (4.6%) |
| 5 | 1 (0.6%) | 5 (2.1%) |
| 6 | 3 (1.7%) | 4 (1.7%) |
| 8 | 0 (0.0%) | 3 (1.3%) |
| 11 | 0 (0.0%) | 1 (0.4%) |
| 19 | 0 (0.0%) | 1 (0.4%) |
| **Sponsor** |  |  |
| Academic | 124 (69.7%) | 190 (80.2%) |
| Pharmaceutical industry | 54 (30.3%) | 47 (19.8%) |

Overall, 3,948 unique outcomes are evaluated in the included studies, including 1,691 from phase 2 trials and 2,257 from later phase trials. We identified 25 generic outcome categories, which are described in table 2. Similar number of outcomes are evaluated in phase 2 (median: 8.5, IQR: 5-13) and later phase (median: 7, IQR: 4-11) trials (supplementary figures 2, 3). While mortality and adverse events are the most frequently assessed outcomes, they are only assessed in 64.6% and 48.4% of all trials, respectively. All remaining outcomes are evaluated in less than half of the trials, highlighting an important heterogeneity in outcomes selection (table 3). Treatment success or failure is only evaluated in 41.6% of phase 2 trials and 44.1% of the later phase trials. Interestingly, the frequency that different outcomes are evaluated as outcomes or as primary outcomes, are very similar for phase 2 and later phase trials.

**Table 2.** Definitions of the generic outcome categories.

| ***Outcome categories*** | **Definitions** |
| --- | --- |
| ***Mortality / Survival*** | Evaluation the survival status. |
| ***Clinical/Physiological*** |  |
| *Treatment success or treatment failure* | A clinical evaluation of whether COVID-19 was successfully treated. Usually a composite endpoint based on one or more of the following: survival, symptoms progression or regression, pyrexia regression, oxygen requirements and/or the requirement for ventilation. We only considered in this category binary outcomes describing criteria either for treatment success or treatment failure. Time-to-treatment success or failure is a measurement instrument that could provide more granular information. |
| *Severity scores* | A quantitative evaluation of disease severity. |
| *Symptoms* | Quantitative or qualitative evaluation of the intensity of one or more symptoms, including but not limited to breathlessness, cough, pyrexia or anosmia. |
| *Oxygenation* | Physiological measures of oxygenation, including oxygen saturation (SatO<sub>2</sub>), the partial pressure of oxygen (PaO<sub>2</sub>) or carbon dioxide (PaCO<sub>2</sub>) |
| *Pulmonary function and physiology* | Measures of pulmonary functions and lung physiology including the forced expiratory volume in 1 second (FEV<sub>1</sub>), forced vital capacity (FVC), respiratory muscle strength or the lung compliance. |
| *Viral detection and load* | Polymerase chain reaction (PCR) to evaluate the presence, persistence and/or load of the severe acute respiratory syndrome coronavirus-2 (SARS-CoV2). |
| *Viral antibodies* | Detection of the presence and titres of antibodies against SARS-CoV2. |
| *Radiological outcomes* | Radiological progression in chest x-ray (CXR) or computed tomography (CT) of the chest. |
| *Inflammatory biomarkers* | The levels and trajectories of any inflammatory biomarkers, including white blood cells count, lymphocytes, neutrophils, eosinophils, monocytes, CD4+ or CD8+ T cell counts, c-reactive protein, interleukins, tumour necrosis factors, or any other inflammatory biomarkers. |
| *Other biomarkers* | The levels and trajectories of any other biomarkers, including but not limited to kidney function, liver function, haematocrit, coagulation profile, d-dimers, troponin or the brain natriuretic peptide (BNP). |
| *Pharmacokinetics / pharmacodynamics* | Evaluation of the pharmacokinetics and/or pharmacodynamics of the drug interventions (mainly serum levels over time). |
| ***Adverse events*** | Adverse events or grade 3 or more severe adverse events, or serious adverse events, according to the Common Toxicity Criteria for |
|  | Adverse Events (CTCAE). In this category, we also included outcomes evaluating specific adverse events, such as renal failure, liver failure, pulmonary embolism, myocardial infarction or tachyarrhythmias. Treatment discontinuation was also included in this category. |
| **Life impact** | Quantitative assessment of the general well-being of participants. |
| **Resource use** |  |
| *Need for (higher) level of care* | This group of outcomes include the need for (i) hospital admission, (ii) hospital re-admission, (iii) intensive care admission, (iv) invasive ventilation, or need for ECMO. In each category, we also included the composite outcomes consisting of the need for the specific level of care or death. For example: “intensive care admission or death”, as these composite outcomes were developed to account for patients who might have benefitted by the higher level of care but died or patients who were not eligible for the higher level of care due to their baseline clinical status. In studies conducted in the hospital setting, need for hospital admission at a specific follow-up timepoint, refers to the proportion of patients who remain inpatients at that timepoint. Similarly, for studies conducted in the ICU stay and the need for ICU admission. |
| *Duration of stay in a specific level of care* | This group of outcomes include length of (i) hospital stay, (ii) intensive care admission, or (iii) mechanical ventilation. The end date was often defined as the last day of stay in a specific level of care, or the last day that the stay was indicated (to account for cases when patients are medically optimized for hospital discharge but remain at hospital for social or other reasons). |
| *Need for supplemental oxygen or NIV* | An assessment of the need for supplemental oxygen, the required oxygen flow or modality of delivery (e.g. oxygen, continuous positive airways pressure [CPAP], bilevel positive airway pressure [BiPAP], or high flow nasal oxygen). |
| *Duration of supplemental oxygen or NIV* | An evaluation of the duration of supplemental oxygen needs. |
| *Need for other organ support* | This category included the need for (a) vasopressors and (b) need for renal replacement therapy. |
| **Other outcomes** | In this category we summarized outcomes that were reported in less than 10 of all eligible trials. These included changes in activities of daily living, quality of life, pharmacodynamics and pharmacokinetics, drug compliance, feasibility outcomes, use of antibiotics or other drugs, emergency room visits or use of other healthcare resources, the need for prone positioning, need for transfusion and discharge destinations. |

**Table 3a.** Frequency that outcome measures are reported in RCTs on the management of COVID-19. Outcomes evaluated in <10 RCTs were grouped as “Other outcomes”. Time to treatment success or failure is a measurement instrument of the outcome treatment success or failure. However, it is reported separately here, as it provides more granular information. A. Grouped in phase 2 and later phase trials. B. Grouped by recruitment setting (community, hospital, ICU). NIV: Non-invasive ventilation. * Continued need of hospital/critical care admission, at a specific timepoint.

| Outcome category | Phase 2 trials (n = 178) |  | Later phase trials (n = 237) |  |
| --- | --- | --- | --- | --- |
|  | Any outcome | Primary outcome | Any outcome | Primary outcome |
| **Mortality / survival** | 115 (64.6%) | 24 (13.5%) | 153 (64.6%) | 32 (13.5%) |
| **Clinical / physiological outcomes** |  |  |  |  |
| Treatment success or treatment failure | 70 (39.3%) | 31 (17.4%) | 103 (43.5%) | 69 (29.1%) |
| Success | 55 (30.9%) | 19 (10.7%) | 88 (37.1%) | 54 (22.8%) |
| Failure | 23 (12.9%) | 12 (6.7%) | 31 (13.1%) | 14 (5.9%) |
| Subgroup: Time to treatment success or treatment failure | 37 (20.2%) | 12 (6.7%) | 62 (26.2%) | 36 (15.2%) |
| Success | 30 (16.9%) | 9 (5.1%) | 59 (24.9%) | 33 (13.9%) |
| Failure | 8 (4.5%) | 3 (1.7%) | 11 (4.6%) | 3 (1.3%) |
| Severity scores | 76 (42.7%) | 21 (11.8%) | 93 (39.2%) | 25 (10.5%) |
| Symptoms | 43 (24.2%) | 5 (2.8%) | 60 (25.3%) | 7 (3.0%) |
| Oxygenation | 63 (35.4%) | 22 (12.4%) | 72 (30.4%) | 23 (9.7%) |
| Pulmonary function / physiology | 12 (6.7%) | 3 (1.7%) | 9 (3.8%) | 1 (0.4%) |
| Viral detection and load | 59 (33.1%) | 20 (11.2%) | 97 (40.9%) | 36 (15.2%) |
| Viral antibodies | 17 (9.6%) | 0 (0.0%) | 8 (3.4%) | 2 (0.8%) |
| Radiological outcomes | 25 (14.0%) | 3 (1.7%) | 25 (10.5%) | 9 (3.8%) |
| Inflammatory biomarkers | 69 (38.8%) | 7 (3.9%) | 66 (27.8%) | 9 (3.8%) |
| Other biomarkers | 47 (26.4%) | 4 (2.2%) | 51 (21.5%) | 2 (0.8%) |
| Pharmacokinetics / pharmacodynamics | 10 (5.6%) | 0 (0.0%) | 5 (2.1%) | 0 (0.0%) |
| **Adverse events** | 95 (53.4%) | 18 (10.1%) | 121 (51.1%) | 8 (3.4%) |
| **Life impact** | 3 (1.7%) | 1 (0.6%) | 10 (4.2%) | 0 (0.0%) |
| **Resource use** |  |  |  |  |
| Hospital admission | 21 (11.8%) | 9 (5.1%) | 30 (12.7%) | 18 (7.6%) |
| Hospital re-admission | 6 (3.4%) | 1 (0.6 %) | 3 (1.3%) | 0 (0.0%) |
| Length of hospital stay | 70 (39.3%) | 5 (2.8%) | 103 (43.5%) | 7 (3.0%) |
| ICU admission | 35 (19.7%) | 6 (3.4%) | 38 (16.0%) | 2 (0.8%) |
| Length of ICU stay | 42 (23.6%) | 0 (0.0%) | 49 (20.7%) | 3 (1.3%) |
| Need for supplemental oxygen or NIV | 31 (17.4%) | 12 (6.7%) | 44 (18.6%) | 3 (1.3%) |
| Duration of supplemental oxygen or NIV | 40 (22.5%) | 2 (1.1%) | 39 (16.5%) | 1 (0.4%) |
| Need for invasive ventilation | 62 (34.8%) | 16 (9.0%) | 87 (36.7%) | 27 (11.4%) |
| Duration of invasive ventilation | 65 (36.5%) | 9 (5.1%) | 68 (28.7%) | 9 (3.8%) |
| Need for vasopressors | 11 (6.2%) | 0 (0.0%) | 10 (4.2%) | 0 (0.0%) |
| Need for renal replacement therapy | 6 (3.4%) | 0 (0.0%) | 7 (3.0%) | 0 (0.0%) |
| **Other outcomes** | 31 (17.4%) | 2 (1.1%) | 42 (17.7%) | 5 (2.1%) |

**Table 3b.** Frequency that outcome measures are reported in RCTs on the management of COVID-19. Outcomes evaluated in <10 RCTs were grouped as “Other outcomes”. Time to treatment success or failure is a measurement instrument of the outcome treatment success or failure. However, it is reported separately here, as it provides more granular information. A. Grouped in phase 2 and later phase trials. B. Grouped by recruitment setting (community, hospital, ICU). NIV: Non-invasive ventilation. * Continued need of hospital/critical care admission, at a specific timepoint.

| Outcome category | Community (n = 63) |  | Hospital (n = 304) |  | ICU (n = 27) |  |
| --- | --- | --- | --- | --- | --- | --- |
|  | Any outcome | Primary outcome | Any outcome | Primary outcome | Any outcome | Primary outcome |
| **Mortality / survival** | 19 (30.2%) | 3 (4.8%) | 216 (71.6%) | 44 (14.5%) | 24 (88.9%) | 8 (29.6%) |
| **Clinical / Physiological Outcomes** |  |  |  |  |  |  |
| Treatment success or treatment failure | 25 (39.7%) | 15 (23.8%) | 140 (46.2%) | 81 (26.6%) | 2 (7.4%) | 0 (0.0%) |
| Success | 16 (25.4%) | 7 (11.1%) | 121 (39.8%) | 63 (20.7%) | 1 (3.7%) | 0 (0.0%) |
| Failure | 12 (19.0%) | 8 (12.7%) | 41 (13.5%) | 17 (5.6%) | 1 (3.7%) | 0 (0.0%) |
| Subgroup: Time to treatment success or treatment failure | 12 (19.0%) | 5 (7.9%) | 83 (27.3%) | 40 (13.2%) | 0 (0.0%) | 0 (0.0%) |
| Success | 7 (11.1%) | 3 (4.8%) | 79 (26.0%) | 37 (12.2%) | 0 (0.0%) | 0 (0.0%) |
| Failure | 4 (6.3%) | 2 (3.2%) | 13 (4.3) | 3 (0.9%) | 0 (0.0%) | 0 (0.0%) |
| Severity scores | 16 (25.4%) | 5 (7.9%) | 136 (44.7%) | 40 (13.2%) | 12 (44.4%) | 1 (3.7%) |
| Symptoms | 31 (49.2%) | 4 (6.3%) | 61 (20.1%) | 7 (2.3%) | 2 (7.4%) | 0 (0.0%) |
| Oxygenation | 6 (9.5%) | 2 (3.2%) | 110 (36.2%) | 35 (11.5%) | 15 (55.6%) | 7 (25.9%) |
| Pulmonary function / physiology | 1 (1.6%) | 1 (1.6%) | 12 (3.9%) | 1 (0.3%) | 5 (18.6%) | 0 (0.0%) |
| Viral detection and load | 35 (55.6%) | 18 (28.6%) | 107 (35.2%) | 34 (11.1%) | 7 (25.9%) | 0 (0.0%) |
| Viral Antibodies | 4 (6.3%) | 0 (0.0%) | 19 (6.3%) | 2 (0.7%) | 1 (3.7%) | 0 (0.0%) |
| Radiological outcomes | 4 (6.3%) | 3 (4.8%) | 40 (13.2%) | 8 (2.6%) | 3 (11.1%) | 0 (0.0%) |
| Inflammatory biomarkers | 6 (9.5%) | 1 (1.6%) | 114 (37.5%) | 14 (4.6%) | 11 (40.7%) | 1 (3.7%) |
| Other biomarkers | 4 (6.3%) | 0 (0.0%) | 79 (26.0%) | 5 (1.6%) | 10 (37.0%) | 0 (0.0%) |
| Pharmacokinetics / Pharmacodynamics | 2 (3.2%) | 0 (0.0%) | 13 (4.3%) | 0 (0.0%) | 0 (0.0%) | 0 (0.0%) |
| **Adverse events** | 25 (39.7%) | 3 (4.8%) | 166 (54.6%) | 21 (6.9%) | 18 (66.7%) | 2 (7.4%) |
| **Life Impact** | 0 (0.0%) | 0 (0.0%) | 7 (2.3%) | 0 (0.0%) | 3 (11.1%) | 0 (0.0%) |
| **Resource Use** |  |  |  |  |  |  |
| Hospital admission | 32 (50.8%) | 21 (33.3%) | 15 (4.9%)* | 4 (1.3%)* | 1 (3.7%)* | 0 (0.0%)* |
| Hospital re-admission | 0 (0.0%) | 0 (0.0%) | 9 (3%) | 1 (0.3%) | 0 (0.0%) | 0 (0.0%) |
| Length of hospital stay | 9 (14.3%) | 1 (1.6%) | 152 (50%) | 11 (36.2%) | 10 (37.0%) | 1 (3.7%) |
| ICU admission | 8 (12.7%) | 0 (0.0%) | 61 (20.1%)* | 8 (2.6%)* | 2 (7.4%)* | 0 (0.0%)* |
| Length of ICU stay | 5 (7.9%) | 1 (1.6%) | 70 (23.0%) | 1 (0.3%) | 14 (51.9%) | 1 (3.7%) |
| Need for supplemental oxygen or NIV | 4 (6.3%) | 0 (0.0%) | 68 (22.4%) | 13 (4.3%) | 1 (3.7%) | 0 (0.0%) |
| Duration of supplemental oxygen or NIV | 3 (4.8%) | 0 (0.0%) | 70 (23.0%) | 3 (0.9%) | 3 (11.1%) | 0 (0.0%) |
| Need for invasive ventilation | 7 (11.1%) | 2 (3.2%) | 130 (42.8%) | 34 (11.2%) | 6 (22.2%) | 4 (14.8%) |
| Duration of invasive ventilation | 5 (7.9%) | 1 (1.6%) | 106 (34.9%) | 10 (3.3%) | 19 (70.4%) | 7 (25.9%) |
| Need for vasopressors | 0 (0.0%) | 0 (0.0%) | 18 (5.9%) | 0 (0.0%) | 2 (7.4%) | 0 (0.0%) |
| Need for renal replacement therapy | 0 (0.0%) | 0 (0.0%) | 10 (3.3%) | 0 (0.0%) | 3 (11.1%) | 0 (0.0%) |
| **Other outcomes** | 13 (20.6%) | 3 (4.8%) | 44 (14.5%) | 5 (1.6%) | 8 (29.6%) | 0 (0.0%) |

**Figure 2.**
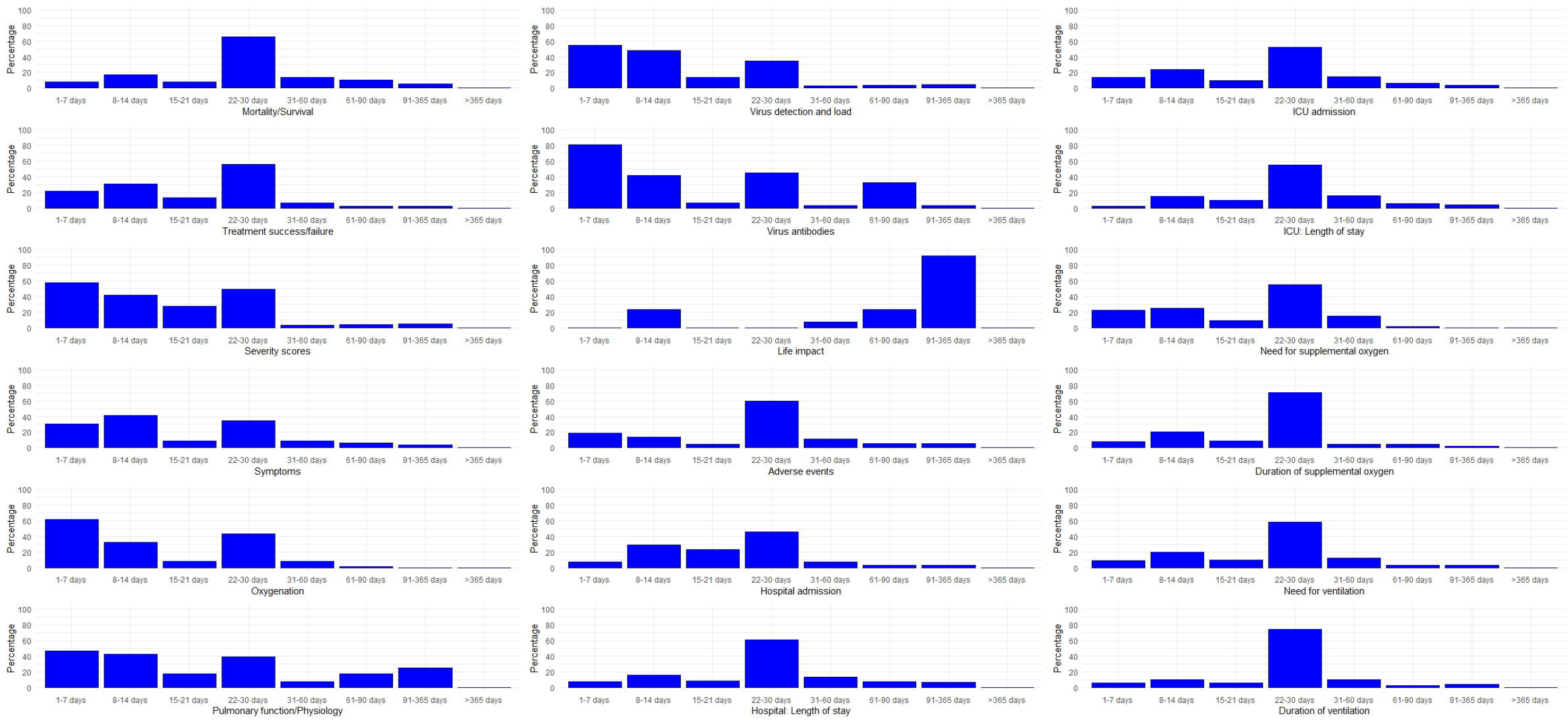
Planned follow-up timepoints for the most frequently evaluated outcomes. All timepoints described in each of the included trials were included in this figure. Presented as a percentage of the outcomes of the same category.

The most frequently reported outcomes among studies conducted in a community setting (thus recruiting less severely ill patients), were viral detection or load (55.6%), and the need for hospital admission (50.8%). In contrast, the most frequently evaluated outcomes in studies recruiting patients with more severe COVID-19, were mortality and adverse events, which were evaluated in 71.6% and 50.3% of studies recruiting hospitalized patients, and in 88.9% and 66.7% of those recruiting critically ill patients, respectively (table 3).

### Outcome measurement instruments

### Mortality/ survival

Mortality is assessed by 284 outcomes. All-cause mortality is evaluated in all but six trials measuring mortality. When mortality was not further described, we presumed it referred to all-cause mortality. Time to death is assessed in 16 trials, while cause-specific mortality is evaluated in six trials, mainly focusing on SARS-CoV2 mortality, but also including mortality due to pulmonary or due to cardiovascular complications.

### Clinical outcomes

#### (Time to) Treatment success or treatment failure

Treatment success or the time to treatment success was evaluated by 220 outcomes. Ordinal scales describing different levels of COVID-19 severity are used for assessing treatment success in 113 (51.4%) of these outcomes. Most scales are very similar to the most frequently used WHO scale, which is a 9-point ordinal scale (from 0 to 8), with each point describing a worse clinical status: no clinical or virological evidence of infection; ambulatory without limitation in daily activities; ambulatory but with limitation in daily activities; hospitalized but not requiring oxygen therapy; hospitalized and requiring oxygen therapy via a face mask or nasal prongs; requiring non-invasive ventilation (NIV) or high-flow oxygen; requiring intubation and mechanical ventilation; requiring ventilation and additional organ support, including vasopressors, renal replacement therapy or extra-corporeal membrane oxygenation (ECMO); deceased^16^. Treatment success is defined as an improvement in ordinal scales such as the WHO clinical progression scale by 2 points or 1 point in 57.5% and 24.8% of all outcomes using the scale to evaluate treatment success, while in the remaining outcomes, no specific threshold is provided. Complete resolution of the symptoms and signs of COVID-19 (clinical recovery) is used as a measure of treatment success in 51/220 (23.2%) outcomes and clinical improvement in 38/220 (17.3%) outcomes. The definition of complete resolution varies. Often, no further information is provided. In the remaining cases, it is defined as a composite outcome including several of the following components: complete resolution of breathlessness, tachypnoea, hypoxia, desaturation, cough, anosmia, myalgia, fever, or of oxygen requirements; a negative COVID-19 PCR; hospital discharge; or radiological resolution. A definition of clinical improvement as an outcome is also frequently lacking. In the remaining cases, it is defined as an improvement in several of the previously listed components. Improvement is either based on prespecified thresholds, or on a subjective clinicians’ judgement. Finally, 14 outcomes (6.4%), use specific thresholds (0, ≤2 or ≤4) of the National Early Warning Score (NEWS or NEWS-2) to define treatment success.

Treatment failure, or time to treatment failure is evaluated by 76 outcomes. In most cases (40/76, 52.6%), treatment failure is defined as a composite outcome consisting of several components with clear thresholds, such as: death, need for ICU admission, need for invasive ventilation, need for other organ support (e.g. vasopressors or renal replacement therapy), need for NIV, need for supplemental oxygen, a deterioration in oxygenation, need for hospital admission or re-admission or emergency visit, ventricular tachyarrhythmia. Ordinal clinical severity scales such as the WHO scale are used to define treatment failure in 16/76 (21.1%) outcomes, while the need for rescue therapy is used in 9/76 (11.8%) outcomes. The definitions are less well defined in the remaining 11 (14.5%) outcomes, which do not provide any criteria or state treatment failure will be based on the clinician’s judgement of deterioration in the clinical condition of the patient.

#### Severity scores

Standardized scores are used to evaluate disease severity and progression in 277 outcomes. In this category we included outcomes presenting mean/median scores or change from baseline in a score. Outcomes describing predefined scores thresholds for treatment success or failure were classified in the previous category. Ordinal disease severity scales (such as the WHO scale) are the most frequently used score (144/277 outcomes, 51.2%), followed by the Sequential Organ Failure Assessment (SOFA) Score^18^, a validated score for describing the severity of organ dysfunction (54/277 outcomes, 19.5%), and the National Early Warning Score (NEWS or NEWS2, 36/277 outcomes, 13.0%), a tool developed by the Royal College of Physicians, that quantifies clinical severity and deterioration by evaluating six physiological measures, namely respiratory rate, oxygen saturation, temperature, systolic blood pressure, heart rate and the level of consciousness^19^. Acute Physiology And Chronic Health Evaluation II (APACHE II, 5/277), clinical sign score (5/277), Pneumonia Severity Index (PSI, 3/277), BRESCIA-COVID, Murray score, Sepsis Induced Coagulopathy, Small Identification Test, SMART-COP score, and the Vienna Vaccine Safety Initiative (ViVI) disease severity score are used less often.

#### Symptoms

188 outcomes focus on symptoms, which are either assessed using visual analogue scales, or validated instruments. Composite scores evaluating several symptoms, including breathlessness, cough, sputum production, pyrexia, anosmia, myalgia, headache, or gastrointestinal symptoms, are evaluated in 40 outcomes (21.3%). Four composite outcomes specifically assess respiratory symptoms (2.2%). Each of the remaining outcomes focus on a specific symptom. These include fever (72/188, 38.3%), breathlessness (18, 9.6%), cough (12, 6.4%), and less often anxiety, depressive symptoms, anosmia, cognitive dysfunction, nausea, insomnia or fatigue. In this category we also included the assessment of heart rate (8, 4.3%) or blood pressure (5, 2.7%).

### Physiological outcomes

#### Oxygenation

Oxygenation parameters are evaluated by 215 outcomes. The need for supplementary oxygen or ventilation were summarized in separate outcome categories. Oxygenation is evaluated using the partial pressure of oxygen (PaO^2^), fraction of inspired oxygen (FiO^2^), oxygen saturation (SatO^2^), or respiratory rate. In this category we also included measurements of the partial pressure of carbon dioxide (PaCO^2^) and pH, which are only rarely evaluated as outcomes. Oxygenation is often measured as the PaO^2^ or SatO^2^ corrected for FiO^2^ (95/215, 44.2%).

#### Pulmonary function and physiology

Twenty-eight outcomes assessed pulmonary function or physiology. There is significant heterogeneity in this domain, with different outcomes evaluating peak flow rate, forced vital capacity (FVC), the ratio of forced expiratory volume in 1 second (FEV^1^) to FVC, vital capacity, diffusing capacity, lung compliance and respiratory muscle function.

#### Viral detection and load

Viral detection and load is evaluated in 235 outcomes. The vast majority assess virologic clearance by a specific timepoint, or the time until virologic clearance. A small number of outcomes track changes in viral load over time, or differences in the viral detection and load when using different samples (nasal, nasopharyngeal, oropharyngeal swabs or sputum).

#### Viral antibodies

The development of antibodies against SARS-CoV2 is assessed in 31 outcomes. Evaluation of specific antibody types (IgA, IgG or IgM) is only described in five trials.

#### Radiological outcomes

Sixty-one outcomes evaluate disease progression radiologically. Definitions of this outcome are inadequate. In most cases, it is broadly stated that the progression, regression, or resolution of the radiological findings are monitored. Details are only provided in six outcomes, which monitor the extent of the lesion as a proportion of the full lung volume, or perform lung densitometry. Development of fibrosis is evaluated in seven outcomes. Computed tomography (CT) is used in 21 (34.4%) outcomes, a chest X-ray (CXR) in 8 (13.1%), either a CT or a CXR in 3, either CT or CXR or lung ultrasound in one and nuclear imaging in one outcome. The imaging modality used is not declared in the remaining 28 (45.9%) outcomes.

#### Inflammatory biomarkers

This group includes 321 outcomes, each describing either a single or multiple inflammatory biomarkers. The most frequently evaluated biomarkers are the total white cell count, neutrophils, lymphocytes, eosinophils, monocytes, c-reactive protein, interleukins 1, 6 and 8, followed by other interleukins, procalcitonin, tumour necrosis factors, complement components, lymphocytes subtypes, immunoglobulins, and other inflammatory biomarkers.

#### Other biomarkers

309 outcomes evaluate either a single or multiple non-inflammatory biomarkers. Mostly, these are surrogates for safety or adverse events. The most frequently captured biomarkers are d-dimers, cardiac enzymes, kidney function, liver function, clotting, red blood cells and haemoglobin, followed by a variety of other molecules.

#### Pharmacokinetics/ Pharmacodynamics

We categorized 33 outcomes in this category, mostly evaluating plasma drug concentrations (12/33, 36.4%), but also half time, maximum/minimum observed concentration, time to reach the maximum/minimum observed concentration, area under the plasma concentration-time curve.

### Adverse events

#### Adverse events

448 outcomes focus on adverse events. 108 (24.1%) evaluate any adverse event; either their frequency, or participants experiencing at least one adverse event. 80 (17.9%) outcomes specifically assess serious adverse events, as defined by the Common Terminology Criteria for Adverse Events (CTCAE). Nineteen (4.2%) outcomes focused on drug reactions, 14 (3.1%) on grade 3 or 4 adverse events, as defined by the CTCAE, and 22 (4.9%) assessed the rate of study drugs discontinuation due to adverse events or due to any reason. The remaining outcomes focused on specific adverse events, mostly cardiac (38, 10.3%), secondary infections (37, 10.0%), thrombotic or bleeding events (29, 8.1%), or local administration reactions (13, 3.6%)

### Life impact

Life impact is evaluated by 13 outcomes. The EuroQol 5 Dimensions (EQ-5D) is used in four outcomes, followed by the RAND 36-Item Health Survey (SF-36), which is used in three outcomes. Other instruments include the WHO Disability Assessment Schedule (WHODAS 2.0), the Control, Autonomy and Pleasure (CASP-19) and the Nottingham Health Profile.

### Resources use

#### Need for a (higher) level of care

352 outcomes are included in this category. Need for hospital admission is evaluated by 68 outcomes (19.3%), need for hospital re-admission by 9 (2.6%), need for intensive care admission by 82 (23.4%), need for invasive ventilation by 167 (47.4%), and need for extracorporeal membrane oxygenation (ECMO) by 26 (7.4%; need for ECMO is merged with the outcome need for ventilation in the tables). In studies conducted in the hospital setting, need for hospital admission at a specific follow-up timepoint, refers to the proportion of patients who remain inpatients at that timepoint. Similarly, for studies conducted in the ICU stay and the need for ICU admission. In this category, we also included composite outcomes consisting of one of the above outcomes and mortality (e.g. need for ICU admission or death), as these composite outcomes focus on the need for a higher level of care, while death is added to account for patients who decease before accessing the higher level of care, or those who are not eligible for higher level of care due to their baseline clinical status. Such approaches could be crucial to account for bias, especially in situations such as the COVID-19 pandemic, when hospitals and ICUs are over-burdened and not infrequently unable to accommodate a significant proportion of the patients, leading to the introduction of stricter criteria for triaging patients. Moreover, some outcomes in this category also evaluate time-to-higher level of care (e.g. time-to-hospital admission).

#### Duration of stay in a specific level of care

This category includes 469 outcomes. Of those, 206 (43.9%) focus on the length of hospital stay, 96 (20.5%) on the length of ICU stay, and 167 (35.6%) on the duration of invasive ventilation. Delays in discharging patients who are medically optimized due to social or other reasons could introduce bias in the outcome length of hospital stay. To account for this issue, 11 outcomes are defined as the time to discharge or to a NEWS ≤2, maintained for 24 hours and another outcome as the time until participants are deemed medically optimized for discharge by a clinician.

#### Need for supplemental oxygen or NIV

This category includes 105 outcomes evaluating the need for supplemental oxygen or NIV in any setting. Most evaluate the need for supplemental oxygen administration at specific follow-up timepoints; 34 (32.4%) outcomes assess the need for NIV (including continuous positive airway pressure [CPAP] or bilevel positive airway pressure [BiPAP]), and 21 (20.0%) the need for high-flow oxygen. One outcome evaluates the need for domiciliary oxygen after hospital discharge. On several occasions, the need for supplemental oxygen was defined on the basis of ordinal clinical severity scales (such as the WHO scale), as the proportion of participants grouped in milder categories, not requiring supplemental oxygen.

#### Duration of supplemental oxygen or NIV

95 outcomes were included in this category. Twelve (12.6%) evaluate the duration of NIV, and seven (7.4%) the duration of high-flow oxygen.

#### Need for other organ support

Forty-four outcomes focus on the need for organ support other than invasive ventilation, including 26 (59.1%) assessing the need for vasopressors, and 18 (40.9%) for renal replacement therapy.

#### Other outcomes

Here, we grouped 145 outcomes that could not be categorized in the previous categories and were evaluated in <10 RCTs each. Need for concurrent treatments are assessed in 22 outcomes, including 7 that specifically focus on the administration of antibiotics. Exercise capacity is assessed by 13 outcomes (mostly using the 6-minutes walking test), COVID-19 transmission by 9, resource requirements and costs by 8 outcomes. Other outcomes include the use of prone positioning, ability to perform activities of daily living, incidence and progression of cytokine storm syndrome, resilience, lost workdays and discharge destinations.

### Study follow-up

Planned follow-up for all included studies varies from less than a week, to over a year (figure 1, supplementary figure 4). However, in most cases, it does not exceed one month (263/415 63.4%). Follow-up exceeds four months only in 50 (12.0%) studies and one year only in one. Follow-up plans do not differ between phase 2 and later phase trials, where it are limited to one month or less in 105/178 (59.0%) and in 158/237 (66.7%) trials, respectively. Longer-term follow-up, exceeding 4 months, is planned for 163 outcomes (figure 2, supplementary figure 5), evaluating mortality (16 outcomes), adverse events (15), life impact (12), severity scores (12), length of hospital stay (11), viral detection and load (11), inflammatory biomarkers (7), pulmonary function/physiology (6), need for ventilation (5) and duration of ventilation (5).

## Discussion

In this methodological survey, we analysed the outcomes and outcome measurement instruments used in 415 RCTs evaluating therapeutic interventions for COVID-19. We identified a remarkable heterogeneity in the selection of outcomes, that is not unexpected given that these trials were designed within a few months from the emergence of the new coronavirus strain. More specifically, only 64.6% and 48.4% of the studies evaluate mortality and adverse events, respectively, while each of the remaining outcomes is assessed by markedly less than half of the studies.

Variability was also observed in the choice of instruments used to measure different outcomes. Ordinal clinical severity scales, such as the WHO clinical progression scale were consistently used across the included studies to assess treatment success or failure, disease severity and oxygen requirements. Given the acute nature of COVID-19, and significant changes in the clinical status of patients in the course of the disease, such scales can effectively capture disease progression, especially in more severe presentations. Most of these scales follow the structure of the WHO scale, removing scale points for simplicity. Despite sharing a similar structure, these scales group patients differently, limiting interpretability and comparability. The WHO recently introduced a revised 11-point Scale, with increased granularity, and it would be advisable for all studies to align relevant outcomes with this revised scale, to improve interpretability and comparability^12^. To evaluate treatment success or failure, most studies used a 2-point change in the ordinal scale as a threshold, that corresponds to a significant change in the clinical status of the patient and this seems appropriate.

Our study revealed a lack of focus on the long-term sequelae of SARS-CoV2 infection. The planned study follow-up exceeds four months only in 12% of all studies. Moreover, only 13 trials assess life impact beyond the acute phase, while exercise capacity is assessed by 13 trials, and the ability to perform simple daily activities during convalescence in only four trials. Finally, only seven trials stated an intent to explore the development of pulmonary fibrosis. However, persistent symptoms, such as fatigue or breathlessness, and quality of life deficits are detected in many hospitalized patients, two to three months after discharge^20-22^. Moreover, fibrotic changes are detected in about one in three survivors of a hospitalization for COVID-19 infection^23,24^. However, it should be noted that we evaluated RCTs registered by May 2020 and longer-term follow-up may have been planned for newer studies, in view of the emerging data.

While this study did not focus on the analytical approaches used for evaluating outcomes, we observed that several studies described specific approaches to account for the bias introduced by mortality as a competing factor for other outcomes, including the duration of hospital stay, ICU stay and the duration of respiratory support. Several methods were described to account for this bias. Some studies stated the duration of hospital or ICU stay will be censored for deceased participants, while others assessed the days that participants are alive and out of hospital or ICU, instead. Homogenization and detailed description of the analytical approaches in the study protocols, along with the outcomes and outcome measurement instruments are crucial for increasing transparency and comparability. Future methodological studies should address analytical approaches.

Four core outcome sets have already been published, with overlapping but not identical selection of components. The WHO Working Group on the Clinical Characterisation and Management of COVID-19 infection recommends the minimal use of three outcomes: mortality, viral burden and non-mortal clinical outcomes evaluated using the WHO clinical progression scale^12^. WHO also highlighted the need for a longer follow-up, of at least 60 days, to capture disease mortality, which is not adopted by most identified trials. Two other groups prioritized specific outcomes and measurement instruments, all of which were captured in our analysis, but were not necessarily the most frequently used^10,11^. The last core outcome set prioritized broader domains to be addressed, rather than specific outcomes^9^. These domains encompass most outcomes identified in this methodological review. The same group also highlighted the need to evaluate the impact of COVID-19 on patient status and life impact in the longer term. Looking across these core outcome sets, a meta-core outcome set (meta-COS) was identified, only including the two domains that were prioritized by all initiatives (mortality and respiratory support), as the most critical, to be evaluated in all future RCTs in hospitalised patients^25^. Both domains recommended by the meta-COS were evaluated in 205 (49.4%) of the included studies.

In view of the multiple available core outcome sets, the authors of this review believe that outcomes selection for future trials should (i) adhere to the recommendations by the WHO and the meta-COS, and (ii) attempt to address all of the domains proposed by Tong et al, a core outcome set that was informed by consensus of >9,000 participants^9^.

Methodological systematic reviews were conducted as part of the development of three core outcome sets. However, these reviews were almost exclusively based on studies conducted in China. Moreover, two of these reviews included approximately 100 RCT protocols^10,11^, while the WHO document was informed by 1,135 protocols, including both observational and interventional studies^12^. However, the outcomes of RCTs often differ from those selected in observational studies. Our methodological review was based on a globally representative sample of 415 RCTs, it employed more rigorous methodology to assess all outcomes, and it is the first review to evaluate the instruments used to evaluate the different outcomes beyond mortality.

Our study only included clinical trials that were registered until May 2020 and this may be a limitation as trial designs and endpoints may have evolved since then, in view of the emerging knowledge on the nature and outcomes of COVID-19 infection, and the published core outcome sets. Moreover, we only evaluated studies registered with the U.S. National Library of Medicine clinical trials register (ClinicalTrials.gov). However, our extensive, globally representative sample of 415 ongoing RCTs was a major strength of our methodological survey and we strongly believe it was sufficient to capture all relevant outcomes and measurement instruments. Characteristically, after extracting data from approximately 25% of the included trials, it became clear that we reached saturation with regards to the outcome categories, while by the time we extracted approximately 70% of the trials, we also reached saturation with regards to the outcome measurement instruments. Therefore, we are confident that we have not missed important outcomes, although we anticipate that newer outcomes may be introduced in newer trials, in response to our expanding knowledge on COVID-19 natural history and outcomes. Future studies will need to assess the impact of the emerging evidence on the natural history and outcomes of COVID-19 and of the four published core outcome sets and the meta-COS on the selection of outcomes in more recently registered trials. Another limitation of our study is the lack of a prospectively registered protocol. However, we have used rigorous methodology recommended by the COMET Initiative, that we have previously employed in similar methodological systematic reviews^13^.

Overall, this methodological survey reveals significant heterogeneity in the outcome categories and measurement instruments selected by trialists in the management of COVID-19 and highlights the need for greater consistency, to enable decision-makers to compare and contrast studies.

## Data Availability

Not applicable - this is a methodological systematic review

