## supplementary for "Outcomes evaluated in controlled clinical trials on the management of COVID-19: A methodological systematic review"

### Outcomes evaluated in clinical trials on the management of Coronavirus Disease 2019: A methodological survey.

**Contents:**

***Supplemental figure 1.*** *PRISMA flowchart*

***Supplementary figure 2.*** *The association between the number of outcomes reported in each RCT and the study population. Having observed that some trials listed numerous inflammatory or other biomarkers as distinct outcomes, for each RCT we have summarized inflammatory biomarkers as a single outcome and other biomarkers as another single outcome. Supplemental figure 2 is a non-corrected version of this figure. (A) Phase 2 trials, (B) Later phase trials.*

***Supplementary figure 3****. The association between the number of outcomes reported in each RCT and the study population. Non-corrected data. (A) Phase 2 trials, (B) Later phase trials*

***Supplementary figure 4.*** *Duration of follow-up in the included studies. (A) Phase 2 trials, (B) Later phase trials.*

***Supplementary figure 5.*** *Selected follow-up timepoints for the most frequently evaluated outcomes. All evaluation timepoints described in each of the included trials were included in this figure. Presented as a percentage of the outcomes of the same category. (A) Phase 2 trials, (B) Later phase trials.*

***Registration numbers of the included studies***

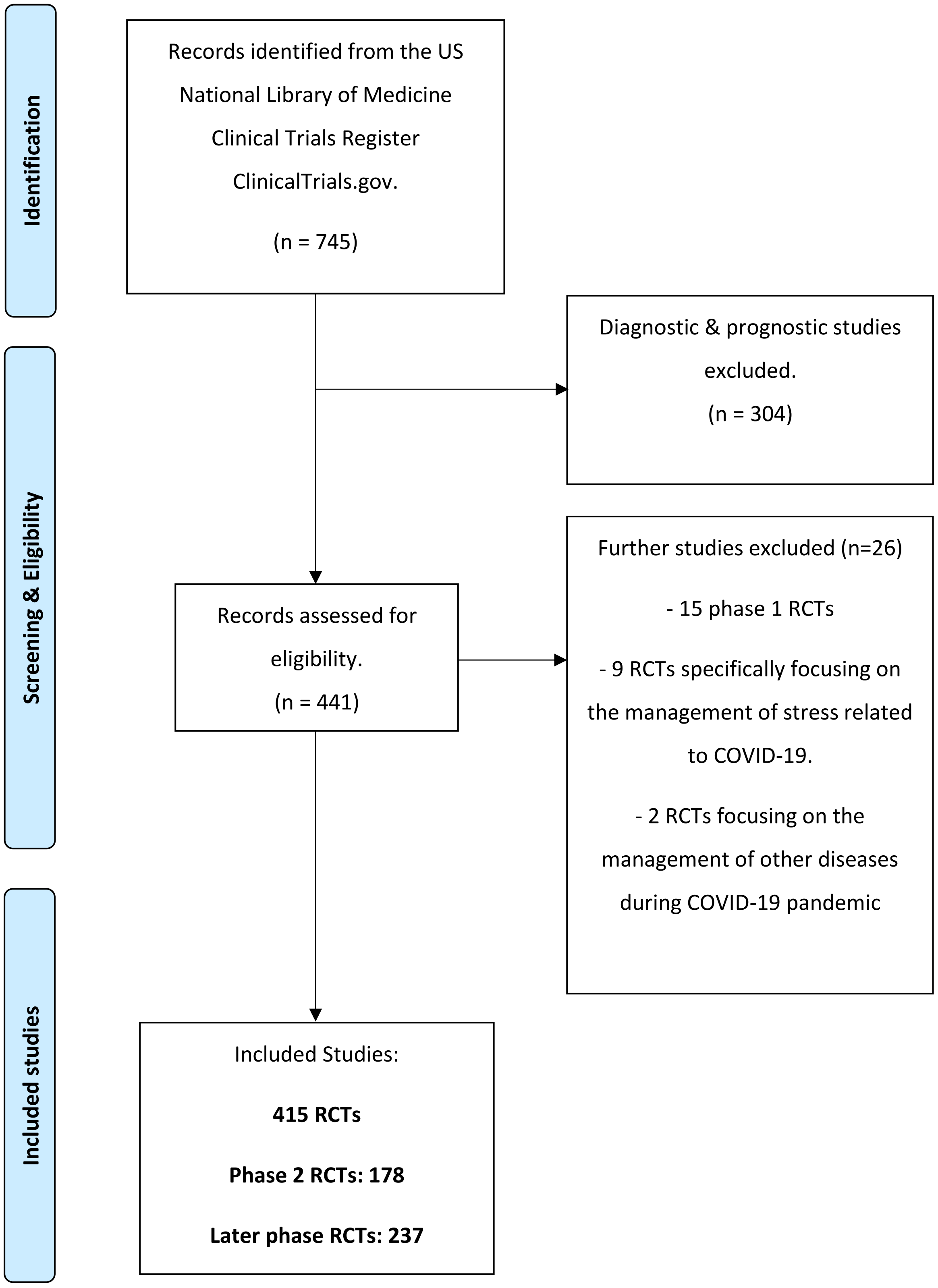


***Supplemental figure 1.*** *PRISMA flowchart*

**A.**

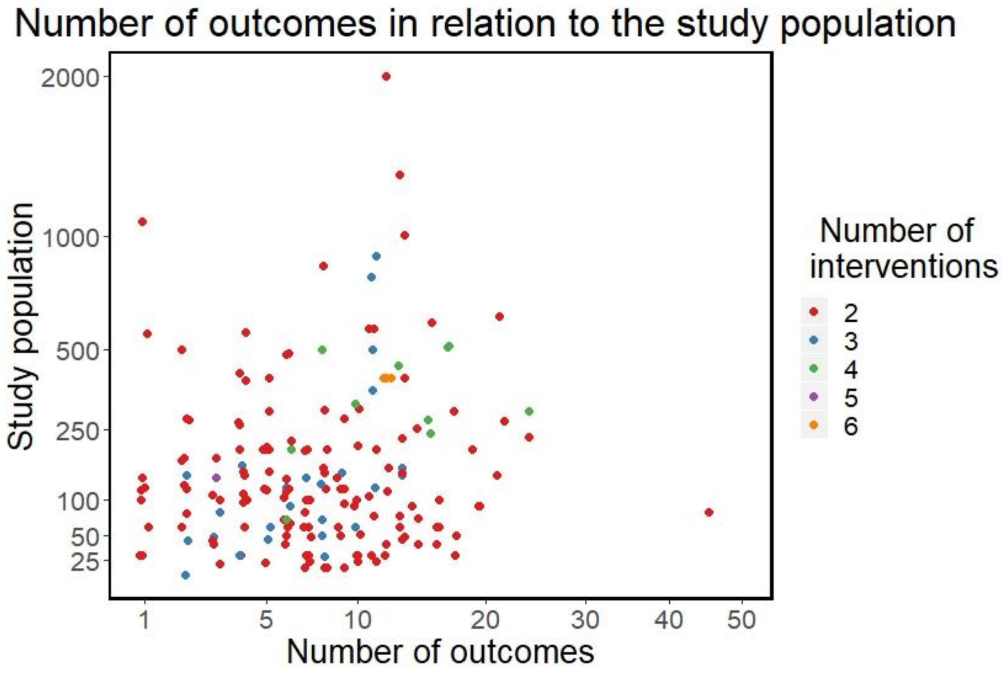


**B.**

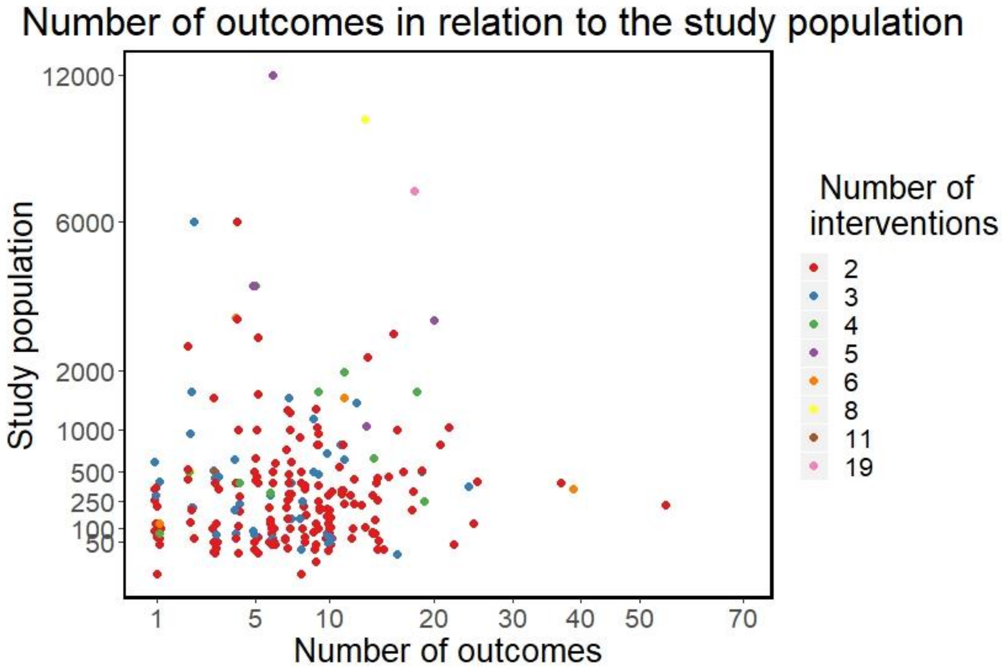


***Supplementary figure 2.*** *The association between the number of outcomes reported in each RCT and the study population. Having observed that some trials listed numerous inflammatory or other biomarkers as distinct outcomes, for each RCT we have summarized inflammatory biomarkers as a single outcome and other biomarkers as another single outcome. Supplemental figure 2 is a non-corrected version of this figure. (A) Phase 2 trials, (B) Later phase trials.*

**A.**

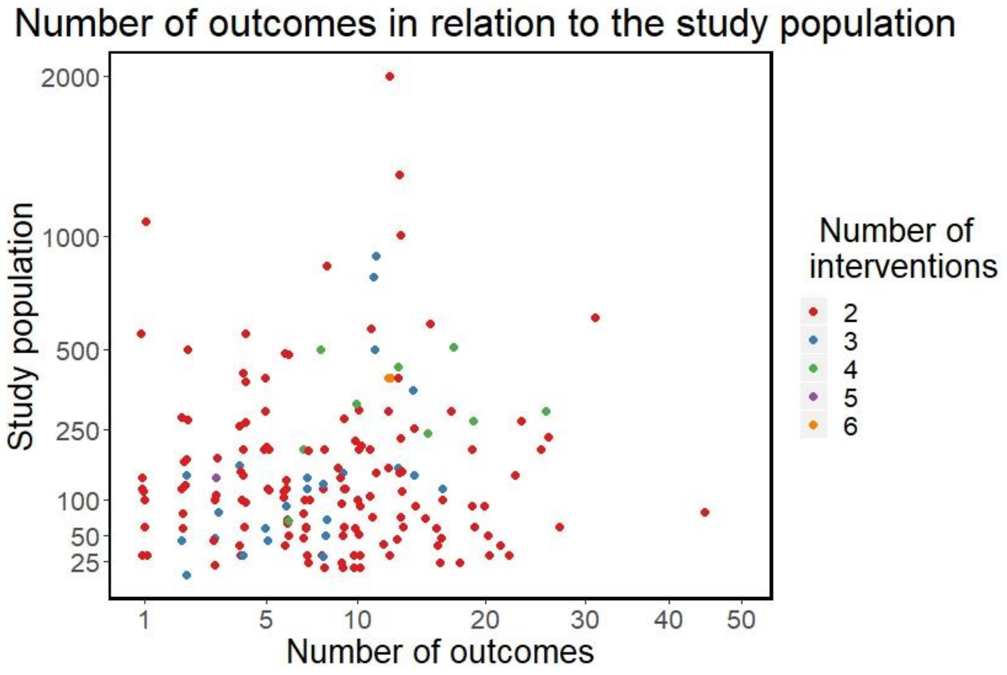


**B.**

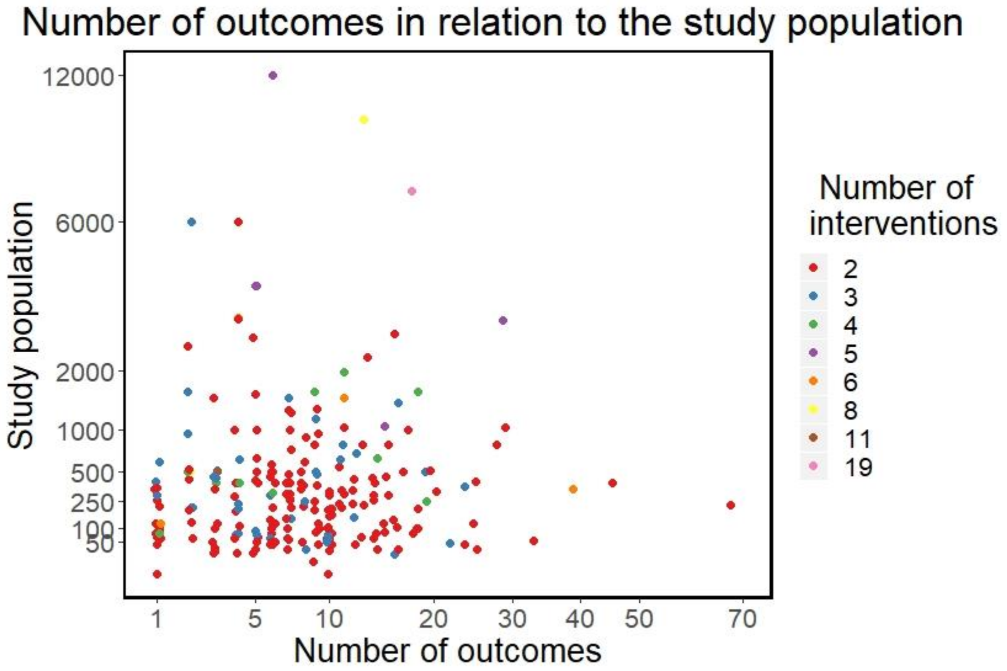


***Supplementary figure 3***. *The association between the number of outcomes reported in each RCT and the study population. Non-corrected data. (A) Phase 2 trials, (B) Later phase trials.*

**A.**

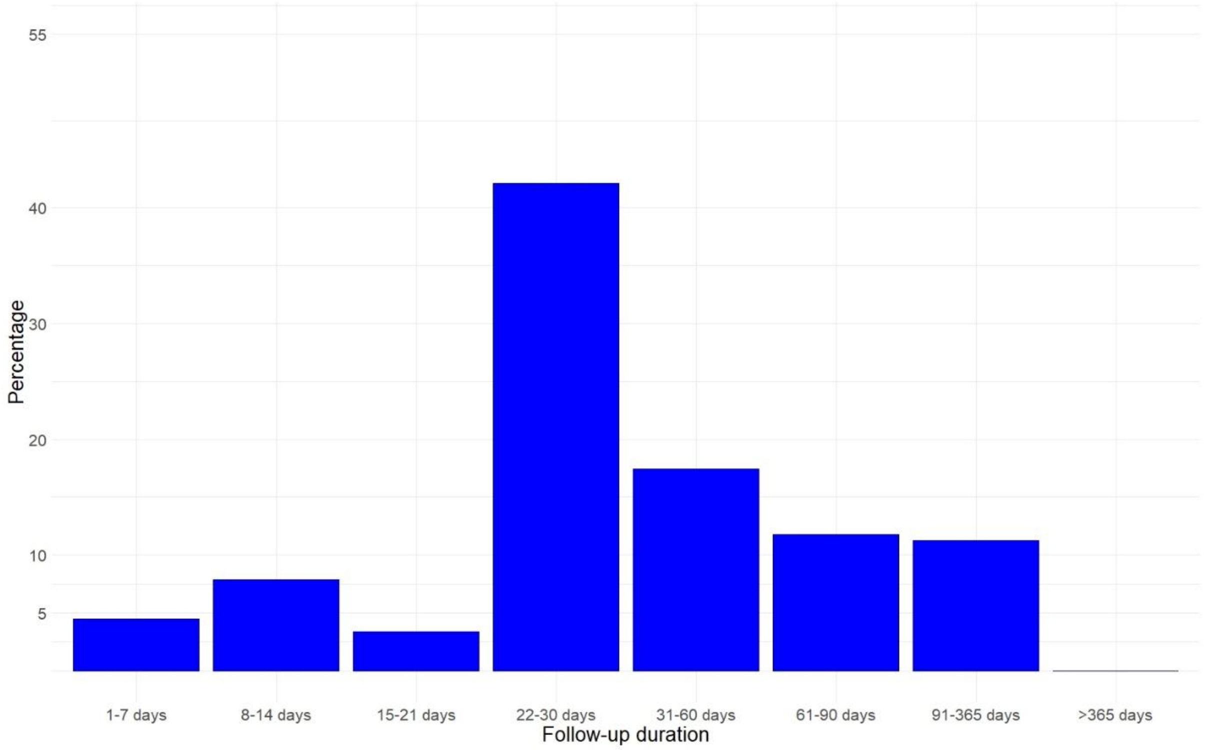


**B.**

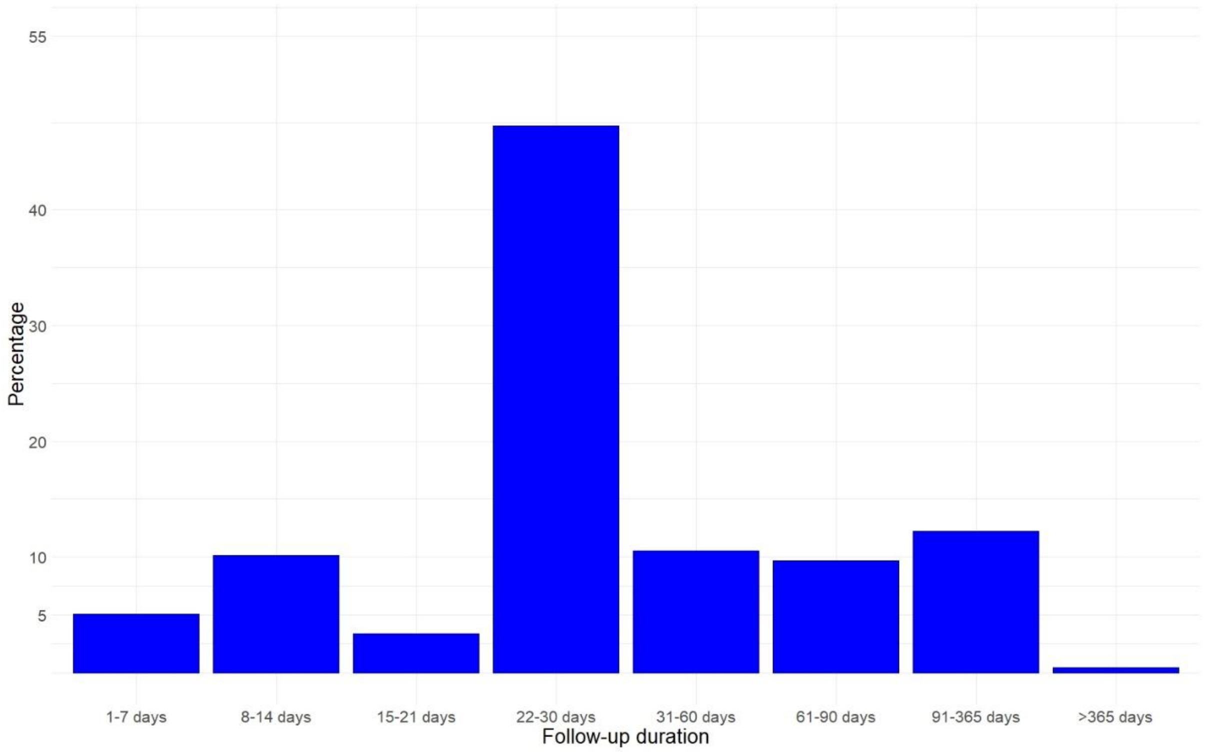


***Supplementary figure 4.*** *Duration of follow-up in the included studies. (A) Phase 2 trials, (B) Later phase trials.*

A.

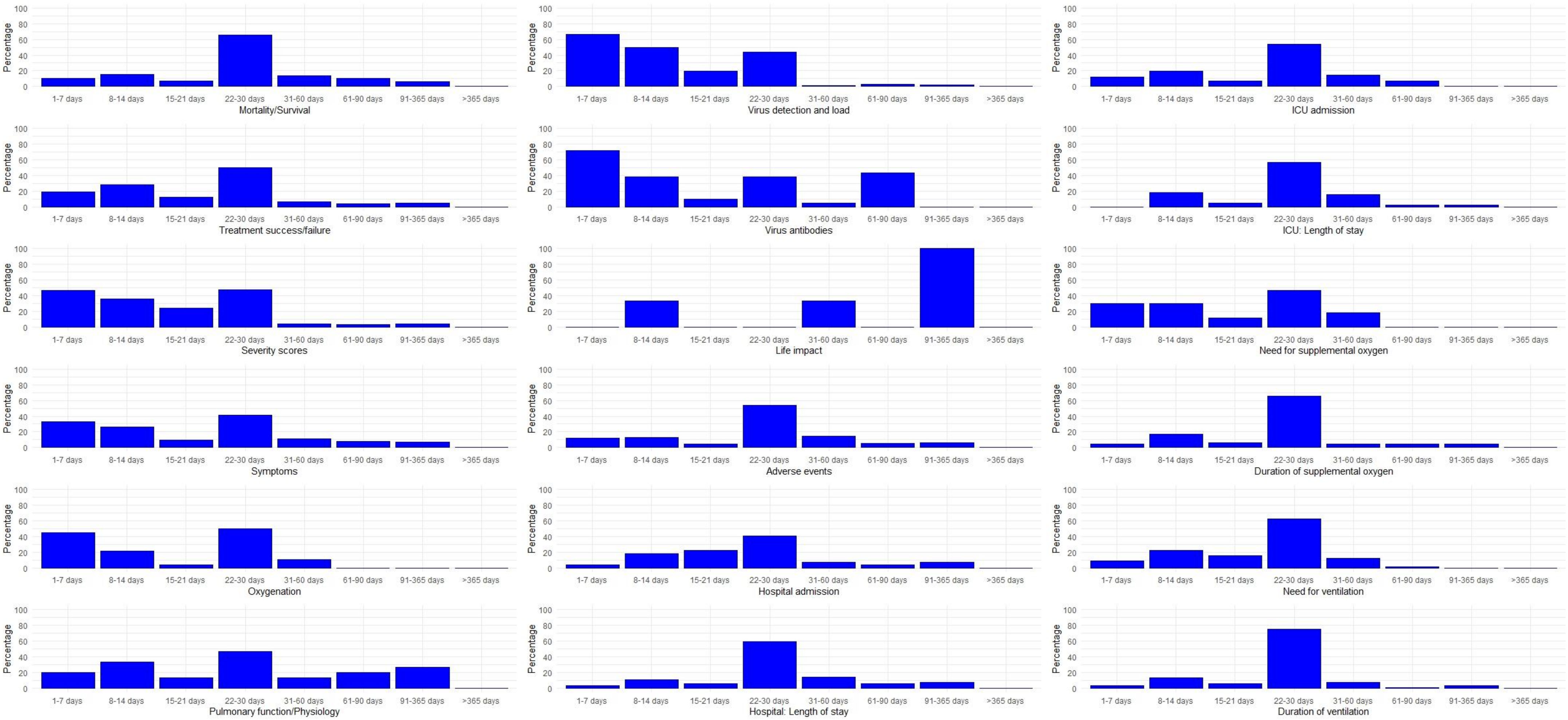

Percentage
1-7 days
8-14 days
15-21 days
22-30 days
31-60 days
61-90 days
91-365 days
>365 days
Mortality/Survival
Virus detection and load
ICU admission
Treatment success/failure
Virus antibodies
ICU: Length of stay
Severity scores
Life impact
Need for supplemental oxygen
Symptoms
Adverse events
Duration of supplemental oxygen
Oxygenation
Hospital admission
Need for ventilation
Pulmonary function/Physiology
Hospital: Length of stay
Duration of ventilation

**B.**

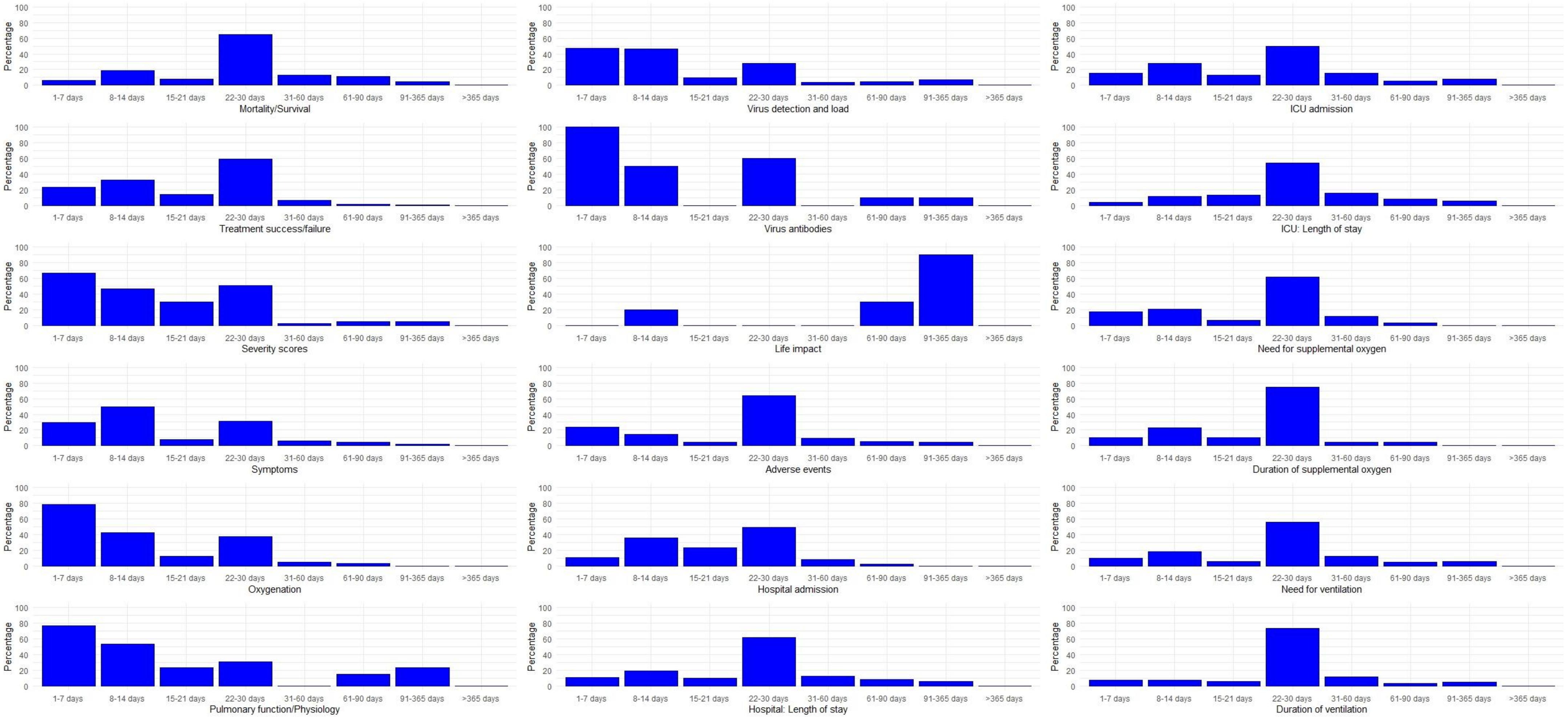


***Supplementary figure 5.*** *Selected follow-up timepoints for the most frequently evaluated outcomes. All evaluation timepoints described in each of the included trials were included in this figure. Presented as a percentage of the outcomes of the same category. (A) Phase 2 trials, (B) Later phase trials.*

**Registration numbers of the included studies**

| NCT Number | N | NCT Number | N | NCT Number | N | NCT Number | N |
|---|---|---|---|---|---|---|---|
| NCT04336904 | 100 | NCT04346147 | 165 | NCT04324528 | 30 | NCT04393038 | 1034 |
| NCT04345445 | 310 | NCT04360876 | 90 | NCT04343651 | 75 | NCT04392141 | 200 |
| NCT04359095 | 1600 | NCT04336332 | 160 | NCT04366908 | 1008 | NCT04405102 | 48 |
| NCT04347915 | 60 | NCT04357444 | 30 | NCT04349618 | 200 | NCT04385043 | 400 |
| NCT04333407 | 3170 | NCT04317040 | 230 | NCT04355962 | 64 | NCT04401579 | 1032 |
| NCT04336462 | 100 | NCT04342897 | 200 | NCT04347239 | 390 | NCT04390594 | 258 |
| NCT04342689 | 1500 | NCT04365231 | 50 | NCT04323800 | 487 | NCT04385940 | 64 |
| NCT04376788 | 15 | NCT04346368 | 20 | NCT04321096 | 580 | NCT04405310 | 80 |
| NCT04333420 | 130 | NCT04371406 | 2770 | NCT04268537 | 120 | NCT04380519 | 372 |
| NCT04360356 | 100 | NCT04288102 | 90 | NCT04332666 | 60 | NCT04382755 | 81 |
| NCT04370262 | 942 | NCT04286503 | 520 | NCT04349098 | 230 | NCT04400890 | 200 |
| NCT04350593 | 900 | NCT04351581 | 215 | NCT04332835 | 80 | NCT04391712 | 20 |
| NCT04372979 | 80 | NCT04351243 | 270 | NCT04361461 | 500 | NCT04394416 | 204 |
| NCT04325633 | 584 | NCT04347512 | 405 | NCT04366271 | 106 | NCT04393311 | 150 |
| NCT04339660 | 30 | NCT04339816 | 240 | NCT04366089 | 152 | NCT04392414 | 60 |
| NCT04362813 | 450 | NCT04347980 | 122 | NCT04373733 | 450 | NCT04398303 | 70 |
| NCT04354389 | 82 | NCT04357808 | 30 | NCT04312009 | 200 | NCT04391127 | 200 |
| NCT04362137 | 402 | NCT04358926 | 30 | NCT04361474 | 120 | NCT04396106 | 180 |
| NCT04359615 | 40 | NCT04293692 | 0 | NCT04315948 | 3100 | NCT04405843 | 400 |
| NCT04359316 | 40 | NCT04362176 | 500 | NCT04311177 | 580 | NCT04381052 | 30 |
| NCT04343768 | 60 | NCT04338828 | 260 | NCT04261426 | 80 | NCT04397562 | 204 |
| NCT04280705 | 800 | NCT04353180 | 45 | NCT04255017 | 400 | NCT04385264 | 800 |
| NCT04329832 | 300 | NCT04371952 | 330 | NCT04254874 | 100 | NCT04385264 | 800 |
| NCT04365257 | 220 | NCT04335305 | 24 | NCT04341935 | 20 | NCT04382586 | 52 |
| NCT04350671 | 40 | NCT04347538 | 90 | NCT04261270 | 60 | NCT04381858 | 500 |
| NCT04350684 | 40 | NCT04372628 | 900 | NCT04342169 | 400 | NCT04379479 | 562 |
| NCT04330586 | 141 | NCT04350320 | 102 | NCT04329195 | 554 | NCT04386616 | 300 |
| NCT04361318 | 100 | NCT04364763 | 252 | NCT04321616 | 700 | NCT04382651 | 120 |
| NCT04361942 | 24 | NCT04344730 | 550 | NCT04311697 | 144 | NCT04393246 | 1407 |
| NCT04315298 | 400 | NCT04341038 | 84 | NCT04357730 | 60 | NCT04390503 | 200 |
| NCT04359953 | 1600 | NCT04328272 | 75 | NCT04367077 | 400 | NCT04394208 | 50 |
| NCT04377620 | 500 | NCT04374487 | 100 | NCT04360096 | 288 | NCT04402866 | 159 |
| NCT04330638 | 342 | NCT04328480 | 2500 | NCT04359810 | 105 | NCT04395170 | 75 |
| NCT04366739 | 40 | NCT04350580 | 138 | NCT03042143 | 75 | NCT04404426 | 100 |
| NCT04369742 | 626 | NCT04323345 | 1000 | NCT04333368 | 40 | NCT04386694 | 30 |
| NCT04363372 | 90 | NCT04366232 | 50 | NCT02735707 | 7100 | NCT04395768 | 200 |
| NCT04326920 | 80 | NCT04342663 | 152 | NCT04348695 | 94 | NCT04402203 | 50 |
| NCT04353284 | 114 | NCT04343001 | 10000 | NCT04347382 | 30 | NCT04391309 | 300 |
| NCT04359277 | 1000 | NCT04356534 | 40 | NCT04358081 | 444 | NCT04389840 | 524 |
| NCT04351763 | 804 | NCT04344288 | 304 | NCT04342650 | 210 | NCT04397718 | 198 |
| NCT04340544 | 2700 | NCT04348383 | 120 | NCT04279197 | 136 | NCT04379076 | 48 |
| NCT04366115 | 126 | NCT04341870 | 27 | NCT04345861 | 7 | NCT04401475 | 510 |
| NCT04366050 | 560 | NCT04352400 | 256 | NCT04376684 | 800 | NCT04401475 | 510 |
| NCT04341675 | 30 | NCT04360824 | 170 | NCT04349592 | 456 | NCT04379271 | 230 |
| NCT04329923 | 400 | NCT04369469 | 270 | NCT04324463 | 4000 | NCT04390061 | 116 |
| NCT04329923 | 400 | NCT04367831 | 100 | NCT04324463 | 4000 | NCT04383535 | 333 |

| | | | | | | | |
|---|---|---|---|---|---|---|---|
| NCT04329923 | 400 | NCT04251767 | 0 | NCT04371393 | 300 | NCT04405921 | 200 |
| NCT04361643 | 120 | NCT04368923 | 60 | NCT04351295 | 40 | NCT04382053 | 120 |
| NCT04355143 | 150 | NCT04348513 | 60 | NCT04363840 | 1080 | NCT04398290 | 30 |
| NCT04333628 | 210 | NCT04257656 | 237 | NCT04351347 | 300 | NCT04392128 | 114 |
| NCT04333628 | 210 | NCT04338126 | 60 | NCT03808922 | 250 | NCT04406532 | 100 |
| NCT04334382 | 1550 | NCT04334850 | 194 | NCT04341493 | 86 | NCT04403646 | 140 |
| NCT04357990 | 81 | NCT04335071 | 100 | NCT04362059 | 24 | NCT04392531 | 120 |
| NCT04335136 | 200 | NCT04348305 | 1000 | NCT04346446 | 29 | NCT04385095 | 400 |
| NCT04362189 | 110 | NCT04362111 | 20 | NCT04365582 | 640 | NCT04390464 | 1167 |
| NCT04330690 | 440 | NCT04355364 | 100 | NCT04363437 | 70 | NCT04381871 | 110 |
| NCT04359511 | 210 | NCT04377503 | 40 | NCT04325906 | 346 | NCT04390139 | 30 |
| NCT04351724 | 500 | NCT04373460 | 1344 | NCT04346628 | 120 | NCT04386447 | 145 |
| NCT04344444 | 600 | NCT04343963 | 436 | NCT04327388 | 409 | NCT04395456 | 144 |
| NCT04344236 | 48 | NCT04349410 | 500 | NCT04344535 | 500 | NCT04401527 | 200 |
| NCT04307693 | 150 | NCT04354428 | 630 | NCT04338906 | 334 | NCT04387760 | 150 |
| NCT04331899 | 120 | NCT04351490 | 3140 | NCT04325893 | 1300 | NCT04393948 | 48 |
| NCT04362332 | 950 | NCT04341415 | 60 | NCT04371367 | 108 | NCT04387240 | 22 |
| NCT04336254 | 20 | NCT04374552 | 140 | NCT04374539 | 116 | NCT04390217 | 120 |
| NCT04332094 | 276 | NCT04365153 | 45 | NCT04251871 | 150 | NCT04397510 | 50 |
| NCT04292899 | 6000 | NCT04356937 | 300 | NCT04361253 | 220 | NCT04390022 | 24 |
| NCT04370782 | 750 | NCT04361032 | 260 | NCT04322123 | 630 | NCT04405570 | 44 |
| NCT04312997 | 100 | NCT04364009 | 240 | NCT04363502 | 30 | NCT04399356 | 100 |
| NCT04377711 | 400 | NCT04353271 | 58 | NCT04322396 | 226 | NCT04399980 | 60 |
| NCT04348409 | 50 | NCT04364737 | 300 | NCT04346693 | 320 | NCT04382040 | 50 |
| NCT04347954 | 45 | NCT04355728 | 24 | NCT04344041 | 260 | NCT04401293 | 308 |
| NCT04360551 | 40 | NCT04366245 | 72 | NCT04321278 | 440 | NCT04379492 | 120 |
| NCT04343989 | 90 | NCT04357457 | 212 | NCT04345289 | 1500 | NCT04389580 | 160 |
| NCT04292730 | 1600 | NCT04333914 | 273 | NCT04358783 | 30 | NCT04384445 | 20 |
| NCT04358549 | 50 | NCT04351191 | 400 | NCT04353037 | 850 | NCT04400929 | 30 |
| NCT04345523 | 278 | NCT04358406 | 60 | NCT04260594 | 380 | NCT04391179 | 80 |
| NCT04346615 | 120 | NCT04326790 | 180 | NCT04326426 | 300 | NCT04405739 | 80 |
| NCT04244591 | 80 | NCT04372082 | 480 | NCT04345406 | 60 | NCT04401150 | 800 |
| NCT04329650 | 200 | NCT04331054 | 436 | NCT04366856 | 500 | NCT04397497 | 50 |
| NCT04331470 | 30 | NCT04344184 | 200 | NCT04338802 | 96 | NCT04402957 | 60 |
| NCT04320615 | 330 | NCT04338698 | 500 | NCT04345887 | 60 | NCT04381377 | 394 |
| NCT04372186 | 379 | NCT04335786 | 651 | NCT04374474 | 75 | NCT04403100 | 1968 |
| NCT04358809 | 480 | NCT04335552 | 500 | NCT04322773 | 200 | NCT04385771 | 80 |
| NCT04273529 | 100 | NCT04357860 | 120 | NCT04345419 | 120 | NCT04381936 | 12000 |
| NCT04374279 | 60 | NCT04351516 | 350 | NCT04347031 | 320 | NCT04402060 | 66 |
| NCT04273581 | 40 | NCT04366063 | 60 | NCT04350281 | 60 | NCT04392778 | 30 |
| NCT04374032 | 120 | NCT04374019 | 240 | NCT04343729 | 416 | NCT04394377 | 600 |
| NCT04363866 | 40 | NCT04356495 | 1057 | NCT04261907 | 160 | NCT04403243 | 70 |
| NCT04342221 | 220 | NCT04346667 | 400 | NCT04264533 | 140 | NCT04402944 | 60 |
| NCT04315896 | 500 | NCT04354441 | 600 | NCT04275388 | 426 | NCT04382846 | 80 |
| NCT04355767 | 206 | NCT04347941 | 200 | NCT04322682 | 6000 | NCT04403555 | 40 |
| NCT04338074 | 100 | NCT04328012 | 4000 | NCT04355052 | 250 | NCT04395807 | 120 |
| NCT04368000 | 60 | NCT04338009 | 152 | NCT04341727 | 500 | NCT04404218 | 480 |
| NCT04331600 | 400 | NCT04310228 | 150 | NCT04346927 | 30 | NCT04380935 | 60 |
| NCT04347174 | 40 | NCT04295551 | 80 | NCT04328467 | 1500 | NCT04404361 | 358 |

| | | | | | | | |
|---|---|---|---|---|---|---|---|
| NCT04363060 | 104 | NCT04365985 | 500 | NCT03852537 | 90 | NCT04389450 | 140 |
| NCT04332107 | 2271 | NCT04273763 | 18 | NCT04367168 | 174 | NCT04395144 | 346 |
| NCT04273646 | 48 | NCT04369794 | 1000 | NCT04276688 | 127 | NCT04396067 | 360 |
| NCT04349241 | 100 | NCT04371107 | 64 | NCT04346940 | 30 | NCT04383717 | 60 |
| NCT04363203 | 300 | NCT04359862 | 50 | NCT03680274 | 800 | NCT04385186 | 60 |
| NCT04252664 | 308 | NCT04324021 | 54 | NCT04308668 | 1309 | NCT04382391 | 20 |
| NCT04341116 | 144 | NCT04334967 | 1250 | NCT04346979 | 50 | NCT04390152 | 40 |
| NCT04375397 | 46 | NCT04298060 | 280 | NCT04401423 | 100 | NCT04380961 | 270 |
| NCT04358068 | 2000 | NCT04332991 | 510 | NCT04406389 | 186 | | |
